# Health and economic impact of introducing norovirus vaccination in England accounting for acute kidney injury: model-based cost-effectiveness analysis

**DOI:** 10.1101/2025.07.31.25332481

**Authors:** Hikaru Bolt, Pratik R Gupte, Laurie A Tomlinson, Rosalind M Eggo, Frank G Sandmann

## Abstract

**Background:** Norovirus vaccines are currently undergoing advanced clinical trials. Acute kidney injury (AKI) is a serious sequelae of norovirus infection. This study evaluated the health and economic impact of implementing a norovirus vaccination programme in England and assessed the contribution of AKI.

**Methods:** We constructed a deterministic age-stratified dynamic-transmission compartmental model. Three single-dose norovirus vaccination strategies were compared to a no-vaccination strategy: targeting children under 5 years of age, targeting adults aged 65 years or older, and targeting both age groups simultaneously. We estimated the impact on preventing primary care attendances, hospitalisations, and mortality from symptomatic norovirus, as well as hospitalisations and mortality due to norovirus-related AKI in adults aged 65 years or older. We evaluated the cost effectiveness over a 10-year time horizon, from a healthcare payer perspective, and discounted costs and quality adjusted life years (QALYs) at 3.5%. We performed probabilistic sensitivity analysis. In one-way sensitivity analysis, we varied the vaccine prices and the proportion of AKI-linked norovirus hospitalisations.

**Results:** A combined vaccination strategy of targeting children and older adults reduced symptomatic infections by 59% (37-84%) and 64% (39-87%) in these age groups, respectively. When including AKI outcomes, all vaccination strategies were cost-effective at £35 per dose and 60% efficacy. At a willingness-to-pay of £20,000 per QALY gained, the combined vaccination strategy had a 96% probability of being the most cost-effective option. Even with a norovirus-related AKI hospitalisation rate as low as 3% among symptomatic norovirus-infected adults, the strategy remained cost-effective. When excluding AKI outcomes, all strategies were not cost-effective.

**Conclusions:** Introducing a norovirus vaccine could be cost-effective in England when accounting for the AKI-related sequelae, which are critical for the health economic evaluation.

## Introduction

Gastrointestinal viruses account for a substantial burden of morbidity and mortality worldwide, and norovirus is one of the leading causes (1,2). There are more than 3 million reported cases of norovirus in the UK annually leading to an estimated economic burden of £300 million on the national health service (3). Outbreaks of norovirus in hospitals cause significant disruption due to control measures and ward closures, especially in winter (3–5). Given the burden, norovirus is a priority pathogen for vaccine research and a number of vaccine candidates have been in development (6–8). With advanced trials of safety and efficacy in progress, evaluating the potential health economic benefits of the vaccines is timely. In the UK, health economic evaluations of vaccines contribute to recommendation decisions by the National Immunisation Technical Advisory Group (NITAG)and the Joint Committee on Vaccination and Immunisation (JCVI).

While norovirus contributes to large outbreaks in hospitals and care homes, in the community only a small proportion of infections are thought to result in hospitalisation and mortality. However, there can be serious sequelae such as acute kidney injury, where the risk is increased more than 40-fold following an episode of gastroenteritis due to hypovolaemia and sepsis (9). Using a population-level model, we previously estimated that 25% (95% uncertainty interval: 15-36%) of symptomatic norovirus infections in individuals aged 65 years or older could be linked to acute kidney injury (AKI) hospitalisations in the winter (10). However, due to limited microbiological confirmation of norovirus, its role in driving AKI hospitalisations has been underrecognized.

Previous studies evaluating the health economic impact of a norovirus vaccine have primarily been considered in the context of the United States (11,12). Both studies demonstrate evidence of cost-effectiveness of different strategies when considering the healthcare payer and societal perspective. No studies have evaluated the potential health economic impact of a norovirus vaccine in the UK, and none have considered the impact of the health-economic burden of norovirus-related AKI admissions.

Therefore, we conducted a study with three objectives. First we assessed the potential epidemiological impact of introducing a norovirus vaccination programme in the UK. Second we evaluated the cost-effectiveness of a potential norovirus vaccination programme from the UK healthcare payer’s perspective following the relevant guidance by JCVI. Third we analysed how norovirus-linked and AKI-related hospitalisations affects the overall cost-effectiveness of the norovirus vaccine.

## Methods

### Vaccination model structure

We used an age-stratified compartmental susceptible-exposed-infected-recovered (SEIR) model for norovirus, which we previously developed and calibrated using both surveillance data and electronic health records (13). We used an age-stratified model in order to characterise heterogeneity in contact patterns and probability of transmission. The dynamic-transmission model structure captures key characteristics of norovirus: a brief incubation period after infection, followed by either symptomatic or asymptomatic progression (Figure 1). Both symptomatic and asymptomatic infections contribute to the force of infection. Individuals recover, however can be infected again in the recovered state, and return to the asymptomatic compartment. Following a period of waning immunity, individuals return to being susceptible.

**Figure 1.**
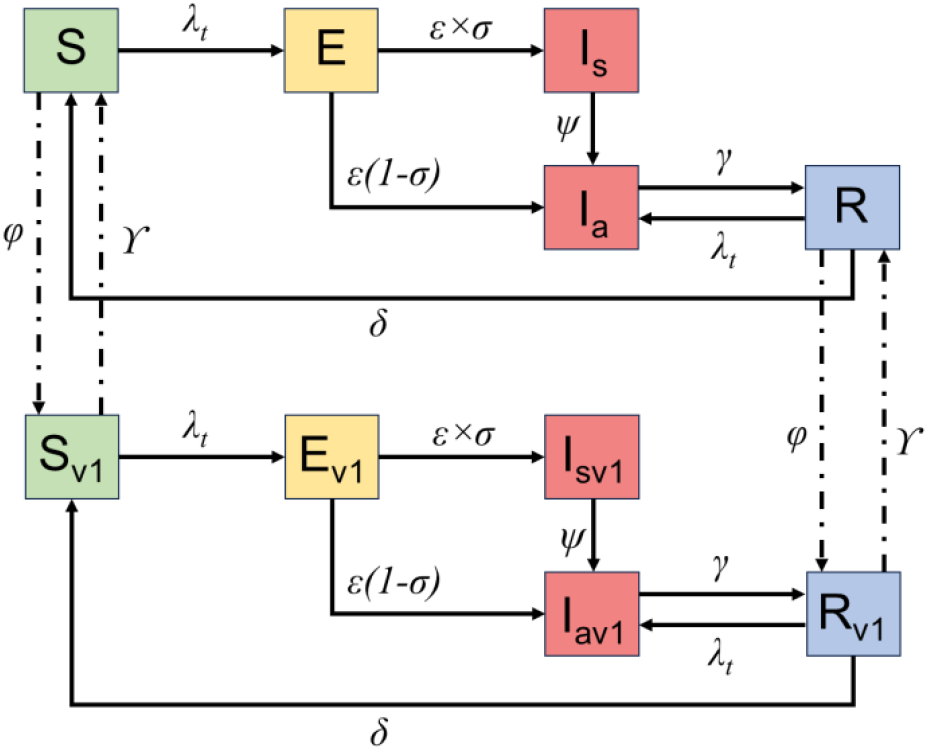
Compartmental model diagram showing epidemic dynamics with vaccination. Compartments represent: S, susceptible (green); E, exposed (yellow); Ia, infectious asymptomatic (red); Is, infectious symptomatic (red); R, recovered (blue). Transition parameters represent: *φ*, vaccination rate; *ϒ*, waning rate of vaccine induced immunity; λ_t_, force of infection; *ε*, incubation period; *σ*, proportion of infections symptomatic; *ψ*, duration of symptoms; *γ*, duration of asymptomatic shedding. Solid arrows denote disease progression (infection, symptom development, recovery) and immunity waning. Vaccinated individuals (v1) follow parallel pathways through corresponding compartments. Dashed lines represent vaccination transitions from unvaccinated to vaccinated states. Births, deaths, age stratification, and aging are modeled but not depicted. Healthcare outcomes of the number of GP attendances, hospitalisations due to norovirus, and hospitalisations due to norovirus linked hospitalisations are calculated using age-group-specific linear factors applied to the number of symptomatic cases (Is) (Supplement S1.4).

We expanded the model to include a one-dose norovirus vaccination schedule. We used a dynamic model in order to capture direct and indirect effects of vaccination between age groups. We conservatively focus on the vaccine’s primary mechanism being symptom reduction rather than preventing infection (6,7). Therefore in our model structure, vaccinated individuals are more likely to experience an asymptomatic episode, if infected. Vaccination initially includes a catch up campaign to achieve coverage within a year in the targeted age group, then shifts to an annual vaccination of people being born or aging into the target groups.

### Epidemiological model parameters and immunisation strategies

The setting of the analysis was England, therefore the model was parameterised based on demographic distribution and contact patterns of the population in England. Estimates for the duration of immunity (mean 8.9 years) and the proportion of symptomatic infection (mean 0.82) were based on data fitted to electronic health data in England (Supplement S1.2) (10). Due to the uncertainty of the duration of vaccine induced immunity, we took a moderately conservative estimate of 2 years of immunity based on previous modelling studies (14,15); this was further explored in a sensitivity analysis ranging from 6 months to 8.9 years. We derived the duration of symptoms, asymptomatic shedding, incubation period, relative infectiousness during asymptomatic period, probability of infection (by age group), and seasonal forcing from a combination of parameter inference and previous studies (Supplement S1.2).

We compared three norovirus vaccination scenarios to no vaccination; vaccination in children aged under 5 years (V1), vaccination in adults aged 65 years and above (V2), and a combination scenario of vaccinating individuals aged under 5 years or 65 years and older (V3). We assumed a coverage of 90% when vaccinating children based on the observed rotavirus childhood immunisation uptake (16). Vaccination coverage in the elderly was estimated to be 75% based on seasonal influenza vaccine uptake. Our baseline analysis assumed a vaccine effectiveness of 60%, meaning 60% of vaccinated individuals progressed to asymptomatic infection, if infected. We additionally conducted a sensitivity analysis for 30% and 90% effectiveness.

### Cost effectiveness analysis

We followed guidance in the UK where only the healthcare provider perspective was considered (17). We modelled a time horizon of 10 years to capture any rebounds in the modelled number of infections. The health outcomes considered in this evaluation were the number of attendances to primary care (general practitioners, GPs) due to norovirus, hospitalisations and death due to norovirus, AKI hospitalisations linked to norovirus infections, and mortality due to norovirus-linked AKI. Hospitalisations and mortality due to AKI were only considered for individuals aged 65 years and older due to it being a syndrome that primarily affects the elderly. We used values from previous studies and fitted values to electronic health data (10) (Supplement S1.4). These outcomes were calculated using age-group-specific linear factors applied to the number of symptomatic cases in each age group which we simulated from the dynamic-transmission model. Age-group-specific linear factors characterised the heterogeneity in risk of suffering more severe health outcomes. We report on outcomes in age groups 0-4, 5-64, and 65+.

We extended the health outcomes from the model to incorporate age stratified quality adjusted life years (QALYs) lost due to primary care attendance for norovirus, hospitalisation for norovirus, and hospitalisation due to norovirus-linked AKI (Supplement S1.5). Health utilities were scaled to one day and then multiplied with the duration of the outcome episode; 12 days for AKI hospitalisations, 5.7 days for the norovirus hospitalisation, and 3 days for a primary care attendance for norovirus (18,19). For norovirus outcomes we considered QALYs lost as the reduction from perfect health that is defined by a health utility of 1.0. For AKI, which is more likely to occur and be recorded in the elderly, QALYs lost were a relative estimate derived from a matched population (19). We also consider life years lost from premature mortality due to norovirus and norovirus-linked AKI.

We discounted life years lost by 3.5% per year and also incorporated a comorbidity adjustment factor on the quality-of-life years lost. We scale down the impact on mortality and quality-of-life years lost due to older individuals and individuals vulnerable to AKI more likely to be frail and have comorbidities compared to the general population (20). All health outcomes were discounted each year by 3.5% after the first year of the vaccine being introduced.

We incorporate costs for GP attendance, activity-weighted AKI hospitalisation costs averaged based on comorbidities, and norovirus hospitalisation costs stratified by children (under 15) and adults (15 and above) (Supplement S1.6). The base year for costs in GBP was 2022/2023; hospitalisation costs and vaccine administration costs were inflated to 2022/2023 costs using the NHS cost inflation index (21–23). We considered vaccine administration costs separately for children under 5 and adults (Supplement S1.6) (23). Vaccine costs were estimated from the reported rotavirus vaccine cost of £35; a range of vaccine costs (£0-500) was further evaluated in a threshold analysis. All costs were discounted each year by 3.5%.

We calculated total discounted costs and QALYs for each scenario to calculate the incremental cost-effectiveness ratio (ICER), and incremental net monetary benefit. These analyses were conducted twice: once evaluating the vaccination scenarios where AKI outcomes were considered, and once in a separate analysis where AKI outcomes were excluded. We carried out a fully incremental analysis of the different vaccination scenarios, including and excluding AKI outcomes.

### Uncertainty analysis

We conducted a one-way sensitivity analysis varying separately all parameter values for the key assumptions on vaccine efficacy, infection induced immunity duration, cost parameters (health care costs, vaccine costs), discounting costs and outcomes, QALYs, and healthcare outcome parameters (e.g. primary care attendance, hospitalisation). In the AKI-included one-way analysis, the proportion of norovirus infections leading to an AKI hospitalisation determined the impact of other variables such as the AKI hospitalisation cost and mortality rate from AKI. We therefore carried out a threshold analysis to explore at which proportion the vaccination strategies stop being cost effective.

Additionally we conducted a probabilistic sensitivity analysis (PSA) sampling a set of all input parameters. We used latin hypercube sampling to efficiently sample 2000 iterations from each distribution. Health outcome parameters were drawn from posterior distributions previously fitted to EHR data following a beta distribution. Costs were assumed to follow a lognormal distribution and health utilities a beta distribution. We present the results of the probability of the vaccination scenarios being cost-effective with cost effectiveness acceptability curves (CEAC) and frontiers (CEAF) to assess the most likely cost effective strategy at different cost-effectiveness thresholds (24). Furthermore, we calculated the expected value of perfect information (EPVI) to quantify the potential value of reducing the decision uncertainty.

## Results

### Epidemiological impact

In the absence of vaccination we estimated an annual incidence of 71 (95% Crl: 46-98) infections in the community per 1000 person years in England over the 10 years. Incidence was highest in the under 5 age group, with an annual incidence of 214 (95% Crl: 156-277) per 1000. Incidence of GP attendance for norovirus was 1.0 (95% Crl: 0.7-1.4) per 1000; age specific incidence for GP attendance was was 9.5 (95% Crl: 6.7 - 12.3) in 0-4 years olds, 3.7 (95% Crl: 2.4-5.1) in 5-14 years olds, 0.1 (95% Crl: 0-0.1) in 15-64 years olds, and 0.1 (95% Crl: 0-0.2) in 65+ year olds per 1000. Annually we estimated 17,135 (9,316-27,300) hospitalisations and 199 (116-298) deaths attributable to norovirus. In addition we estimated 57,734 (27,770-100,297) hospitalisations and 12,765 (3,599-28,509) deaths attributable to acute kidney injury linked to norovirus. To contextualise, if we assumed there were 696,362 all cause AKI hospitalisations annually in the over 65+ over 10 years (10), norovirus linked AKI hospitalisation would account for 8.3% (4.0-14.4%) of all cause AKI hospitalisations.

At 60% efficacy, compared to no vaccination, all three scenarios reduced the number of infections and healthcare outcomes across the simulated period. With 81 million individuals vaccinated, the scenario of vaccinating under 5 and 65+ resulted in 59% (37-84%) and 64% (39-87%) fewer infections in the under 5s and over 65+, respectively (Table 1). This translated to an 64% reduction in norovirus linked AKI hospitalisations in the over 65+; a 5.3% reduction in all cause AKI hospitalisations.

**Table 1.** Estimated total number of individuals vaccinated, infected, and healthcare outcomes (mean and 95% CrI) by vaccination strategy and age groups over 10 years in England.

| Strategy | Age group | Total vaccinated | Infections total | GP attendance | Norovirus hospitalisation | Norovirus mortality | AKI hospitalisation | AKI mortality |
| --- | --- | --- | --- | --- | --- | --- | --- | --- |
| **No vaccination** |  |  |  |  |  |  |  |  |
|  | 0-4 | 0 (0-0) | 7,403,589 (5,330,021-9,262,348) | 329,278 (231,967-424,664) | 24,417 (15,283-34,924) | 46 (33-58) | Not applicable | Not applicable |
|  | 5-14 | 0 (0-0) | 6,155,107 (4,013,639-8,461,206) | 273,692 (180,100-374,632) | 2,462 (1,493-3,667) | 29 (19-40) | Not applicable | Not applicable |
|  | 15-64 | 0 (0-0) | 24,802,299 (15,519,309-34,829,287) | 27,282 (12,897-47,731) | 6,451 (3,689-9,810) | 116 (72-162) | Not applicable | Not applicable |
|  | 65+ | 0 (0-0) | 4,586,029 (2,683,496-6,796,203) | 9,627 (3,640-19,805) | 171,358 (93,166-273,001) | 1,995 (1,162-2,978) | 577,343 (277,705-1,002,972) | 127,658 (35,985-285,089) |
| **V1** |  |  |  |  |  |  |  |  |
|  | 0-4 | 12,757,253 (12,722,958-12,789,221) | 3,184,235 (1,779,125-4,458,134) | 141,628 (77,888-201,239) | 10,500 (5,510-16,219) | 20 (11-28) | Not applicable | Not applicable |
|  | 5-14 | 0 (0-0) | 4,940,340 (2,596,427-7,261,877) | 219,665 (115,543-321,188) | 1,976 (996-3,108) | 23 (12-34) | Not applicable | Not applicable |
|  | 15-64 | 0 (0-0) | 20,926,634 (10,241,585-31,707,407) | 23,017 (9,333-42,360) | 5,444 (2,530-8,851) | 97 (48-147) | Not applicable | Not applicable |
|  | 65+ | 0 (0-0) | 3,854,751 (1,753,002-6,119,769) | 8,091 (2,601-17,282) | 144,000 (65,147-242,622) | 1,677 (764-2,682) | 485,242 (200,489-886,041) | 107,174 (27,731-245,014) |
| **V2** |  |  |  |  |  |  |  |  |
|  | 0-4 | 0 (0-0) | 7,179,001 (5,098,808-9,077,272) | 319,290 (221,198-415,652) | 23,676 (14,507-34,212) | 45 (32-57) | Not applicable | Not applicable |
|  | 5-14 | 0 (0-0) | 5,946,344 (3,812,919-8,259,806) | 264,407 (170,483-365,529) | 2,378 (1,409-3,576) | 28 (18-39) | Not applicable | Not applicable |
|  | 15-64 | 0 (0-0) | 23,602,557 (14,563,869-33,599,647) | 25,962 (12,091-45,756) | 6,139 (3,471-9,417) | 110 (67-156) | Not applicable | Not applicable |
|  | 65+ | 46,323,155 (46,303,091-46,338,661) | 1,985,304 (1,137,606-3,019,462) | 4,168 (1,549-8,677) | 74,175 (39,638-120,003) | 864 (494-1,319) | 249,950 (118,491-441,209) | 55,256 (15,577-122,582) |
| **V3** |  |  |  |  |  |  |  |  |
|  | 0-4 | 12,760,894 (12,727,335-12,792,841) | 3,025,493 (1,591,501-4,301,431) | 134,569 (69,292-194,303) | 9,976 (4,986-15,581) | 19 (10-27) | Not applicable | Not applicable |
|  | 5-14 | 0 (0-0) | 4,711,406 (2,331,899-7,020,010) | 209,482 (104,923-310,721) | 1,884 (887-2,989) | 22 (11-33) | Not applicable | Not applicable |
|  | 15-64 | 0 (0-0) | 19,634,968 (8,972,295-30,221,354) | 21,595 (8,265-39,964) | 5,108 (2,230-8,399) | 91 (42-140) | Not applicable | Not applicable |
|  | 65+ | 46,321,535 (46,301,920-46,337,677) | 1,632,222 (710,401-2,673,847) | 3,426 (1,071-7,531) | 60,967 (25,903-104,887) | 710 (311-1,169) | 205,479 (82,006-386,181) | 45,368 (11,355-105,248) |

V1 strategy demonstrated significant indirect effects, where infections in individuals 5-64 and 65+ were reduced by more than 15% (Figure 2 panel A). V2 strategy primarily demonstrated direct effects in reducing infections, with the most impact in individuals 65+ (Figure 2 panel B). When considering symptomatic cases numbers across different age groups, strategy V3 averts the most infections (Figure 2 panel C). Incidence of symptomatic cases averted per 1000 population demonstrated the same pattern in direct and indirect effects (Supplement S2.2)

**Figure 2.**
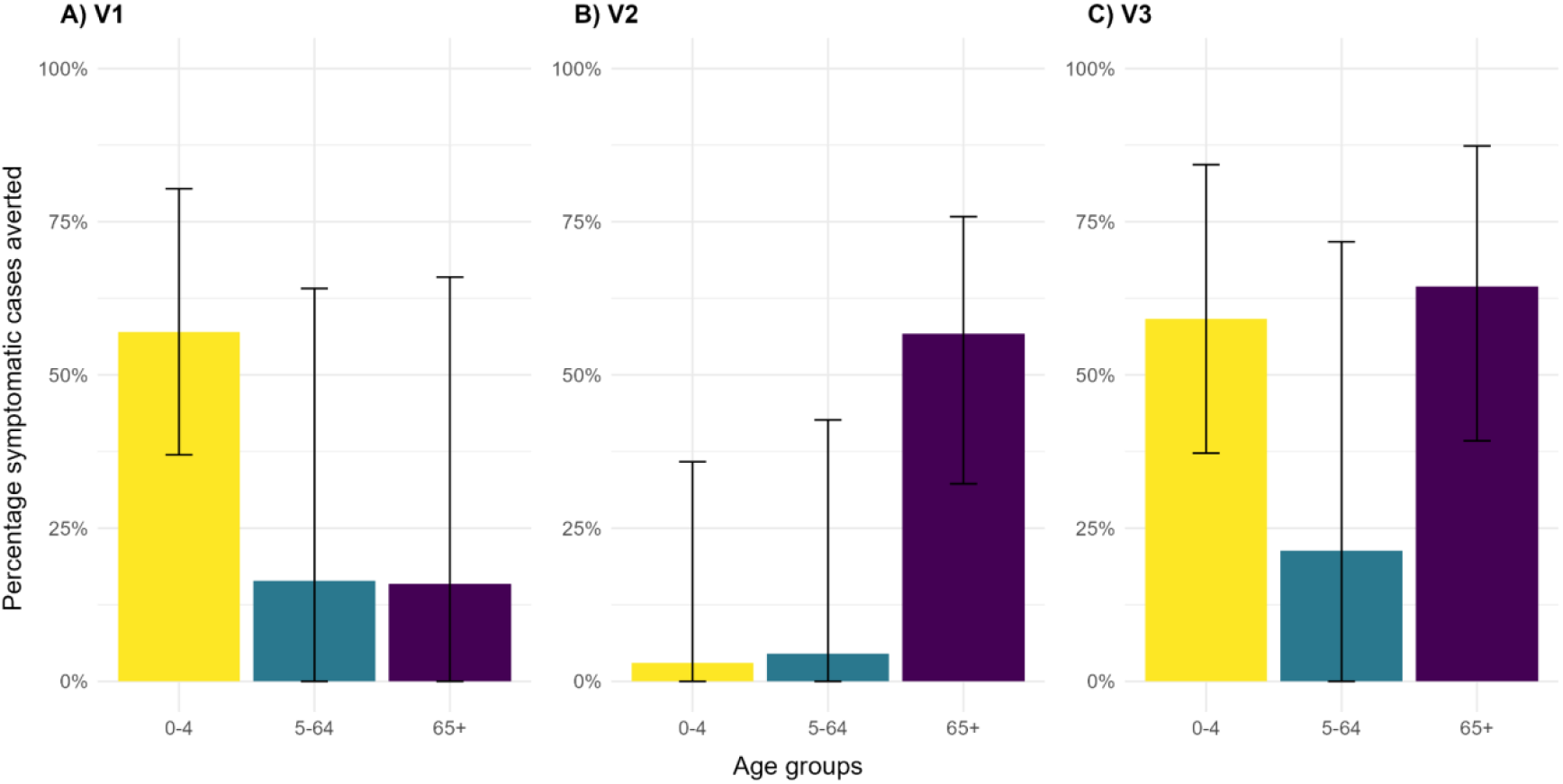
Percentage of symptomatic norovirus infections averted (mean and 95% CrI) per age group by vaccination scenarios in A) V1 B) V2 C) V3. Yellow bars represent percentage reduction in symptomatic cases in children aged 0-4. Blue bars represent percentage reduction in symptomatic cases aged 5-64. Purple bars represent percentage reduction in symptomatic cases aged 65+.

We observed a rebound in infections following a honeymoon period; the timing and size of the rebound differed between age groups and vaccination strategies (Figure 3). In the V1 strategy, we observed a rebound in infections in the 4th season following the introduction of the vaccine in the 5-64 and 65+ age group. With the V2 65+ strategy, significant rebound in infections was not observed across the age groups. Finally in the V3 strategy, rebound in symptomatic infections were observed in the 5-64 age group in the 4th and 5th season following vaccine introduction.

**Figure 3.**
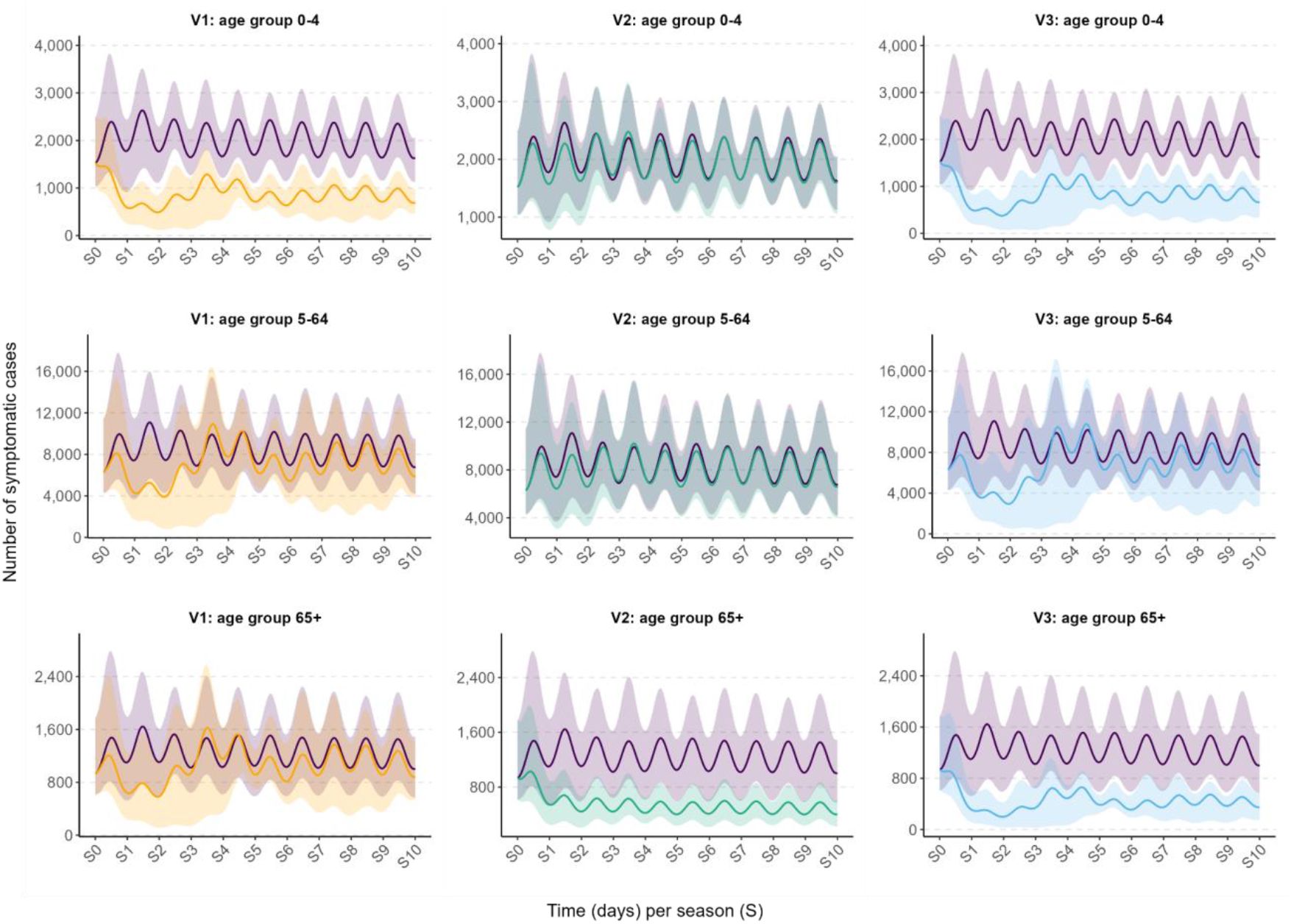
Times series of symptomatic norovirus infections (mean and 95% CrI) by age groups and vaccination scenarios in England over 10 years. Colours denote vaccination strategy: orange is V1, green is V2, and blue is V3. No vaccination is shown in purple. First row plots the number of symptomatic cases in children aged 0-4 years; second row plots the number of symptomatic cases in individuals aged 5-64 years old; and third row plots the number of symptomatic cases in adults aged 65 years and older. Solid line indicates mean and shaded area the 95% CrI.

### Health economic impact

Overall, when considering a vaccine cost of £35, in all scenarios that included acute kidney injury hospitalisations and mortality, all strategies were cost effective at a willingness-to-pay threshold of £20,000 per QALY gained (Table 2). The no vaccination strategy was dominated by all vaccination strategies. Each successive strategy cost more but provided more QALYs gained. We observed that averting AKI mortality contributes the highest QALY gains across the different strategies (Supplement S2.3).

**Table 2.** Cost effectiveness indicators (mean and 95% CrI) by vaccination strategy and inclusion of acute kidney injury outcomes.

| Acute kidney injury outcomes | Strategy | Net cost (£ million ) | Total QALYs gained (million) | Net monetary benefit (£ million) | Average incremental cost-effectiveness ratio (£/QALY) | Incremental cost-effectiveness ratio (£/QALY) |
| --- | --- | --- | --- | --- | --- | --- |
| **Included** |  |  |  |  |  |  |
|  | V1 | 254 (-171 - 539) | 128 (44 - 265) | 2,308 (501 - 5,175) | 1,982 (-1,224 - 9,187) | 1,982 (-1,224 - 9,187) |
|  | V2 | 865 (-890 - 1,981) | 424 (126 - 929) | 7,610 (1,113 - 18,456) | 2,042 (-1,931 - 11,823) | 60 (-4,153 - 6,899) |
|  | V3 | 1,330 (-575 - 2,556) | 487 (151 - 1,048) | 8,412 (1,153 - 20,260) | 2,730 (-1,011 - 12,829) | 688 (-85 - 2,053) |
| **Excluded** |  |  |  |  |  |  |
|  | V1 | 612 (454 - 774) | 4 (3 - 4) | -537 (-699 - -377) | 164,054 (115,638 - 220,868) | 164,054 (115,638 - 220,868) |
|  | V2 | 2,114 (1,557 - 2,727) | 7 (4 - 11) | -1,968 (-2,596 - -1,387) | 289,398 (170,068 - 520,571) | dominated |
|  | V3 | 2,752 (2,141 - 3,397) | 10 (7 - 13) | -2,553 (-3,202 - -1,924) | 276,572 (182,434 - 418,888) | 112,518 (9,814 - 256,016) |

In scenarios that did not include AKI outcomes, all vaccination strategies were not cost effective given that the incremental cost-effectiveness ratios were higher than the willingness-to-pay threshold of £20,000 per QALY. V1 and V3 were non-dominated strategies. V2 65+ was dominated by other strategies providing more value for money. Without AKI outcomes, averting norovirus hospitalisations contributes the highest QALY gains.

### Sensitivity analysis

The top ten parameters which influenced the one-way sensitivity analysis differed whether AKI outcomes were included. With AKI outcomes, ICERs were sensitive to AKI hospitalisation costs, mortality rate from AKI, vaccine cost, duration of vaccine immunity, vaccine efficacy, and the proportion of norovirus infections leading to an AKI hospitalisation in the 65+ age group (Figure 4). With the maximum estimates of these parameters, the ICER would increase by no more than £10,000 per QALY. This kept all strategies below the £20,000 per QALY willingness-to-pay threshold. When excluding AKI, ICERs were sensitive to the vaccine cost, duration of vaccine immunity, vaccine efficacy, cost of hospitalisation due to norovirus, and the proportion of norovirus infections in the 65+ group (Supplement S2.5). Strategy V1 was the least sensitive to changes to these parameters, followed by V3. V2 65+ was the most sensitive with the maximum values of the vaccine cost (£100) and a short vaccine induced immunity duration (6 months).

**Figure 4.**
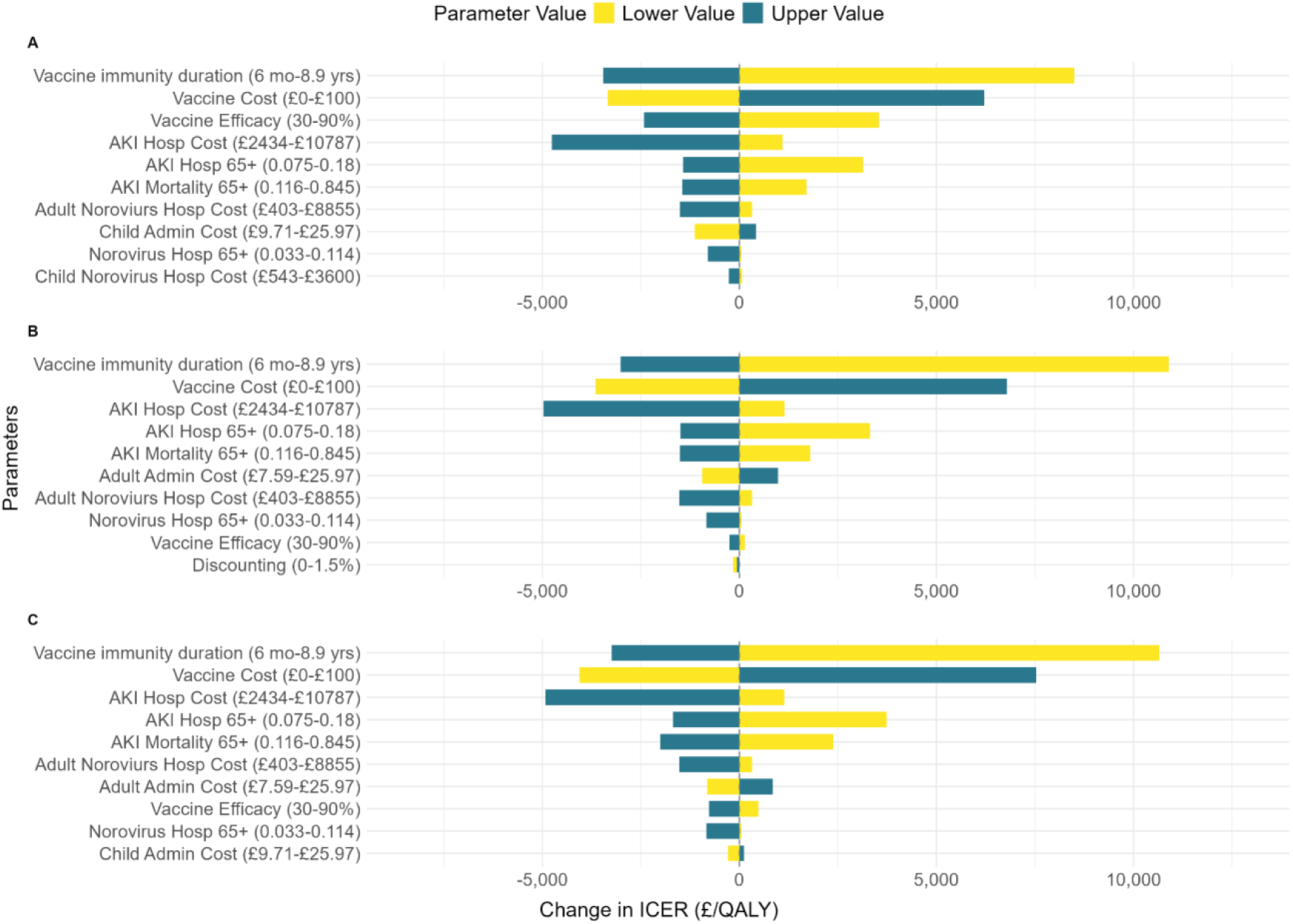
One way sensitivity analysis by vaccination strategy A) V1; average ICER £1,982 per QALY B) V2; average ICER £2,042 per QALY C) V3; average ICER £2,730 per QALY as reported in Table 2. Reported average ICERs vs base case.

For the threshold analysis of the annual proportion of norovirus infections leading to an AKI hospitalisation in the 65+ age group, at a proportion of 0.03 all vaccination strategies remained below a willingness-to-pay threshold of £20,000 per QALY (Figure 5). Strategies were cost saving at an annual proportion of 0.22 for V1, 0.22 for V2 65+, and 0.25 for V3. When examining vaccine cost, all strategies were cost saving were the vaccine to cost less than £20 (Supplement S2.6). All strategies remained below the cost effective threshold if the vaccine cost was less than £190 per dose.

**Figure 5.**
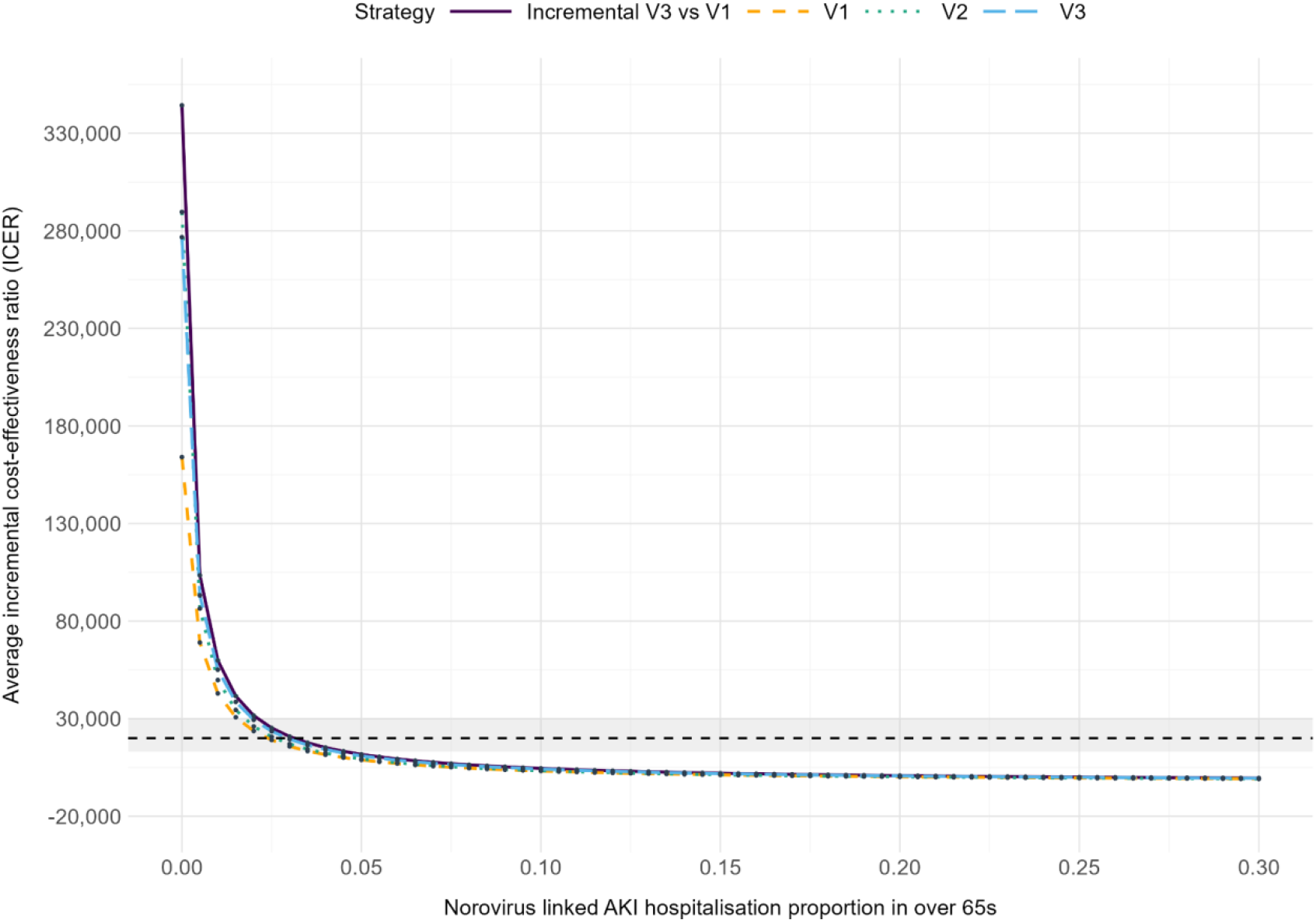
Threshold analysis of the proportion of symptomatic norovirus infections in over 65s with an AKI hospitalisation. Plots of ICERs by vaccination strategy V1, V2, V3, and incremental cost effectiveness ratio V3 vs V1. Dashed line indicates £20,000 ICER. Shaded area indicates ICER £13,000 - £30,000.

The average net monetary benefit at various willingness-to-pay thresholds differed when considering AKI outcomes. With AKI, after a willingness-to-pay threshold of £10,000 per QALY, V3 had the highest net monetary benefit (Supplement S2.7). V3 had the steepest slope which indicated the more cost effective strategy as the willingness to pay increases. Excluding AKI outcomes, strategy V1 had the highest net monetary benefit at lower willingness to pay thresholds. The probabilistic sensitivity analysis, when including AKI outcomes, indicated >99% of samples were cost effective at a threshold of £20,000 per QALY for all vaccination strategies (Figure 6 panel A). Strategies were cost saving in 9% of V1 samples, 11% of V2 samples, and 6% of V3 samples. Without AKI outcomes, <0.2% of samples were cost effective below a threshold of £20,000 per QALY for all vaccination strategies (Figure 6 panel B).

**Figure 6.**
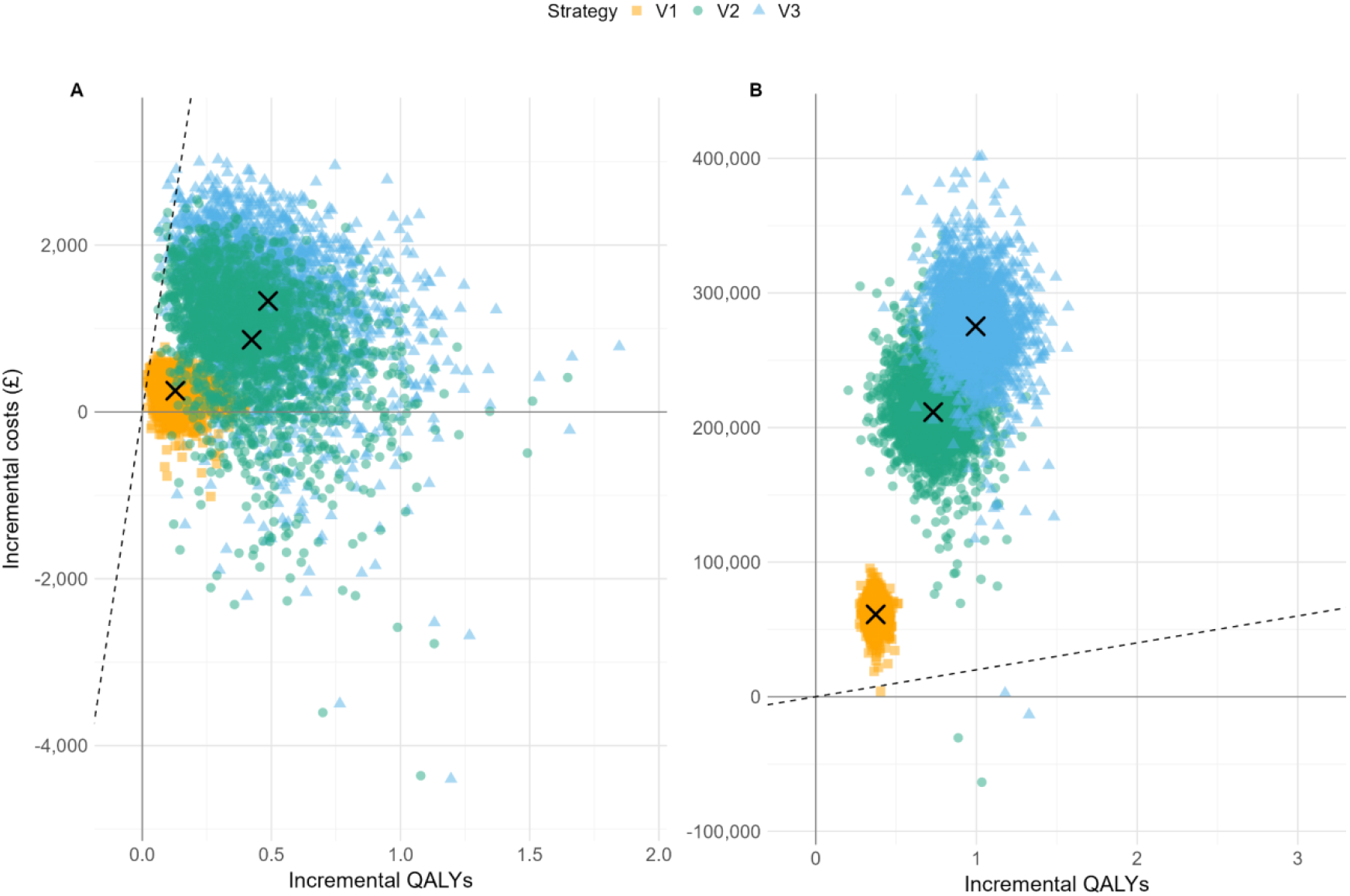
Cost effectiveness plane of probabilistic samples by A) including AKI outcomes, B) excluding AKI outcomes. ‘X’ mark denotes the mean centred around each strategy. Dashed line indicates slope of £20,000 per QALY threshold. Colours denote vaccination strategy: orange is V1, green is V2, and blue is V3.

When comparing the probability of cost-effectiveness between the strategies with AKI included, at a willingness to pay of zero, no vaccination was 84% likely to be the most cost effective strategy (Figure 7 panel A). However, at a willingness-to-pay threshold of £20,000 per QALY and above, strategy V3 had more than a 94% probability of being the most cost-effective strategy. The EVPI peaks at £2,000 per QALY with a value of £389 million (Figure 7 panel E). At a willingness to pay threshold of £20,000 per QALY, EVPI is near zero indicating little decision uncertainty at this point. With AKI excluded, the no vaccination strategy had the highest probability of being the most cost-effective strategy until a willingness-to-pay threshold of £164,000 per QALY (Figure 7 panel F).

**Figure 7.**
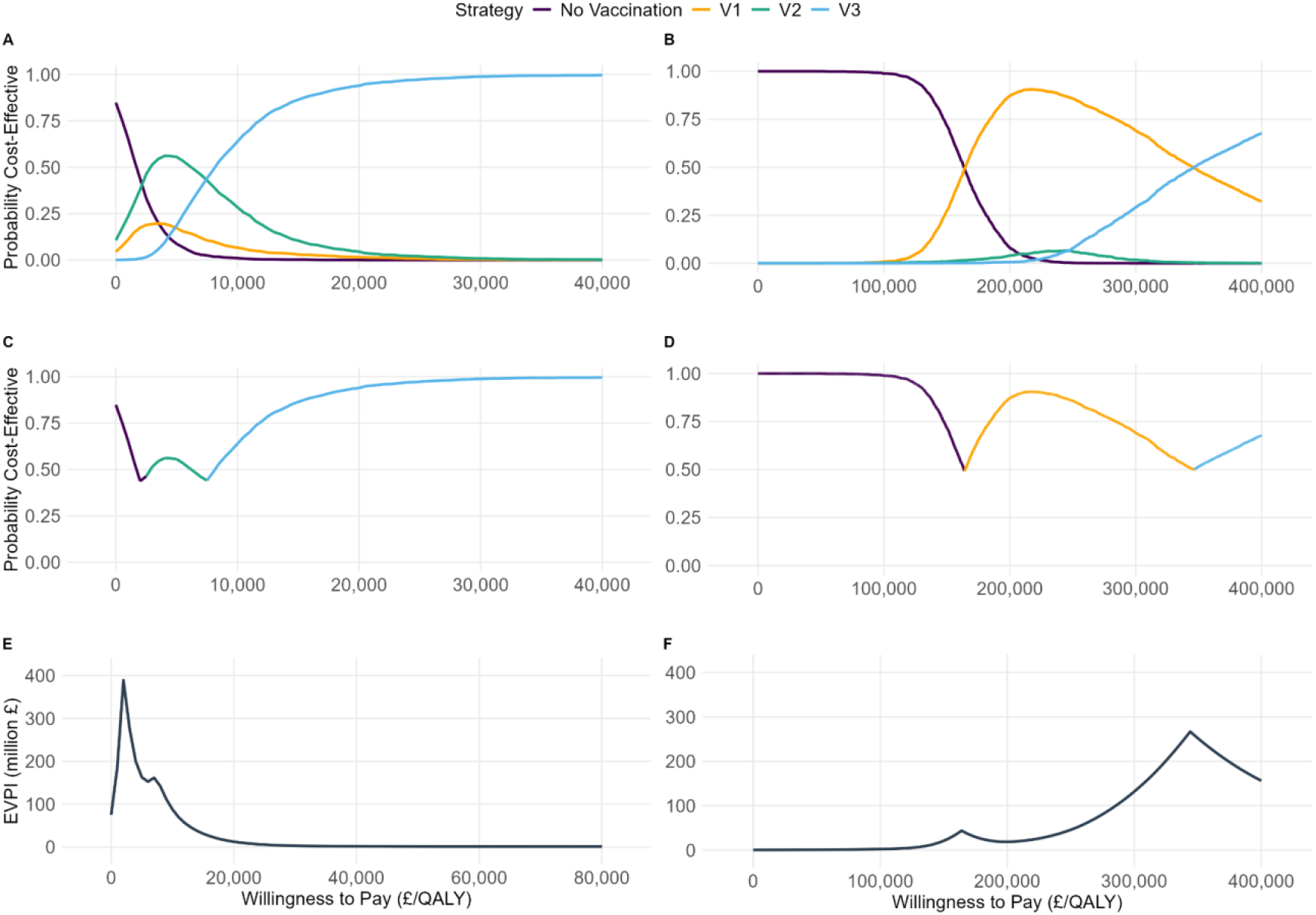
Cost effectiveness acceptability by vaccination strategy A) AKI included and B) AKI excluded. Cost effectiveness acceptability frontier by vaccination strategy C) AKI included and D) AKI excluded. Expected value of perfect information (EVPI) by E) AKI included and F) AKI excluded. Colours denote vaccination strategy: orange is V1, green is V2, and blue is V3. No vaccination is shown in purple.

## Discussion

We conducted a cost effectiveness analysis of norovirus vaccination using a dynamic norovirus transmission model to estimate health economic outcomes. Our results show that when considering norovirus coded healthcare outcomes alone, norovirus vaccination strategies in infants or the elderly would not reach cost effectiveness thresholds. However, when including the AKI outcomes we estimate to be associated with norovirus, all vaccination scenarios demonstrated cost effectiveness. The strategy of vaccinating under 5 and over 65 year olds had the highest potential healthcare impact epidemiological impact and probability of being the most cost effective strategy. At a threshold of £20,000 per QALY, this vaccination strategy would be cost effective at an annual proportion of norovirus-linked AKI as low as 3%.

Previous studies have modelled the impact of norovirus vaccines on community transmission (15,25,26). These studies explored strategies vaccinating children and older age groups, and assumed various durations of vaccine induced immunity. These studies explore a greater range of scenarios, however similar to our findings, vaccinating children provided the greater direct and indirect reduction in infections across all age groups. Targeting older adults provided important direct effects and reduced more severe outcomes, such as hospitalisation and death, but minimal indirect effects.

Cost effectiveness analyses of norovirus vaccination have also previously been conducted in the context of the US (11,12). Steimle et al. developed a dynamic model combined with a decision tree focusing on the impact of vaccinating children in day care settings (12). They demonstrated evidence of cost effectiveness despite a smaller target population. Bartsch et al., used a dynamic model exploring cost effectiveness scenarios of vaccinating children, elderly, and a combination of both, similar to our study(11). However, these studies consider societal perspectives in the cost effectiveness assessment; this is a critical difference to the UK context where the healthcare payer perspective is the primary focus. Therefore, comparisons with these studies are limited.

We observed that the dynamics of norovirus transmission across all age groups were disrupted when vaccinating children under 5. Given the probability of infection was greater from under 5s, we would expect vaccinating under 5s to have greater indirect impacts on the population. The strategy of vaccinating the 65+ age group had an important impact of directly averting the most severe healthcare outcomes, and therefore highest potential for QALY gains. However, to achieve these direct effects, many individuals needed to be vaccinated thus increasing the cost of the strategy. Therefore, indirect effects increased the value of vaccination and potential for QALY gains which was an important factor in the most cost-effective strategies, regardless of the inclusion of AKI as an outcome.

The implication of including AKI outcomes in the cost effectiveness assessment was critical. Inclusion of AKI outcomes made the vaccine cost effective, and shifted the strategy to a combination vaccination strategy, compared to the AKI excluded analysis. Given the impact of AKI on cost effectiveness, it also highlights the importance of the vaccine being available to individuals most vulnerable to AKI, such as those with chronic kidney disease. People with chronic renal conditions were more than 10 times more likely to be admitted to hospital with norovirus than individuals without chronic conditions (27), further highlighting the potential benefit in this group.

There remains some uncertainty in how effective the potential vaccine will be, the duration of vaccine induced immunity, and the cost of the vaccine. The cost effectiveness estimates were shown to be sensitive to this uncertainty. However, we demonstrate even in the most conservative estimates, the burden of AKI was influential to the extent that the absolute changes in the incremental cost-effective ratios remained under a willingness to pay threshold of £20,000 per QALY.

We are first to perform a comprehensive CEA of introducing norovirus vaccination in England that accounts for sequela. The data that we used were the most-recently available and sourced from the literature and national databases. Additionally, using a dynamic transmission model has the advantage of capturing indirect effects. The results of this study can be used to inform national vaccination policy recommendations.

We excluded strain type dynamics in modelling norovirus transmission, and the impact it could have on the efficacy of the vaccine. Norovirus is a genetically diverse virus, and frequent recombination facilitates antigenic shifts and drifts which has important implications in developing natural and vaccine induced immunity (28). Since 2014, there has been a period of stability with the Sydney strain of GII.4 being the dominant strain type in England. However, mixed evidence of within genogroup cross protection and an emergence of GII.17 strain, challenges our model assumptions (29,30). Questions remain whether strain type dynamics influence vaccination strategies sufficiently that an annual vaccination would be required due to reduced longer term effectiveness (26).

This would have important implications on the cost of the vaccination programmes and therefore reframe the potential cost effectiveness of the different strategies. While we assumed a conservative vaccine profile in having 60% efficacy and the primary mode of action reducing symptoms, we maintained this profile through the time horizon which may be unlikely when strain dynamics play an important role in efficacy and immunity. However, if the vaccine demonstrates some level of cross protection across strain types, and prevents infections to some extent, then this would make the vaccine strategies more cost effective. We also did not explore multi-dose strategies, which would further affect cost effectiveness estimates (31).

Our one-way sensitivity analysis excluding AKI outcomes demonstrated uncertainty in many of the parameters that estimate norovirus healthcare outcomes diminishing the ability to assess whether considering norovirus outcomes alone would be cost effective from the provider perspective. We did not include societal costs, which conventionally improve on the cost-effectiveness estimates. We also excluded healthcare opportunity costs, which has previously been shown to be an important cost generated by norovirus outbreaks (3) and would increase the cost effectiveness of vaccination strategies. Ultimately, the inclusion of AKI outcomes minimised the uncertainty and influence of parameters estimating norovirus healthcare outcomes and costs. With the under-recognition of AKI-linked norovirus hospitalisations, we relied on modelled estimates. To improve our certainty and its contribution to the health economic impact, real world data to validate these estimates are important.

## Conclusion

In conclusion, AKI healthcare outcomes were highly influential when evaluating the health economic impact of a norovirus vaccine in a healthcare payer perspective in England. Introducing a norovirus vaccine in the UK would be the most cost effective if vaccinating both under 5s and people aged 65 years and older. Further establishing real-world prevalence of AKI hospitalisations following norovirus infections would bring greater certainty in its contribution to norovirus burden. Future health economic evaluations of norovirus vaccines should consider acute kidney injury outcomes.

## Data Availability

All code and data pertaining to the simulated dynamic model and health economic evaluation are outlined in the manuscript and available at https://github.com/ehr-lshtm/acute_kidney_injury_norovirus_healthecon_evaluation

## Acknowledgements

All code pertaining to the dynamic model and health economic evaluation are available at https://github.com/ehr-lshtm/acute_kidney_injury_norovirus_healthecon_evaluation. The work undertaken has had ethical approval from the London School of Hygiene & Tropical Medicine Observational/Interventions Research Ethics Committee (LSHTM Ethics ref: 22670). HB, FGS, LT, RME contributed to the conception, design, analysis and interpretation of the data for the work. HB contributed to data collection, data curation, and data interpretation. HB, PG contributed to the development of software, and review of code. All authors contributed to the writing, reviewing and editing of the manuscript, approved the final manuscript, and agreed to be accountable for all aspects of the work and publication.

