## Supplemental material for "Health and economic impact of introducing norovirus vaccination in England accounting for acute kidney injury: model-based cost-effectiveness analysis"

### S1. Transmission model and input parameters

#### S1.1 Model equations

*i* denotes age groups: 0-4; 5-14; 15-64; 65+

*î* denotes age groups: 5-14; 15-64; 65+

*k* denotes vaccinating age groups: 0-4; 65+

$$\frac{dS_{0-4}}{dt} = bN + \delta R_{0-4} - \lambda_i S_{0-4} - \varphi_k S_{0-4} + \Upsilon S_{0-4}^{v1};$$

$$\frac{dS_{\hat{\imath}}}{dt} = \delta R_{\hat{\imath}} - \lambda_i S_{\hat{\imath}} - \varphi_k S_{\hat{\imath}} + \Upsilon S_{\hat{\imath}}^{v1} + \tau S_{\hat{\imath}};$$

$$\frac{dE_i}{dt} = \lambda_i S_i - \sigma\varepsilon E_i - \varepsilon(1-\sigma)E_i - dE_i + \tau E_i;$$

$$\frac{dIs_i}{dt} = \sigma\varepsilon E_i - \psi Is_i - dIs_i + \tau Is_i;$$

$$\frac{dIa_i}{dt} = (1-\sigma)\varepsilon E_i + \psi Is_i + \lambda_i R_i - \gamma Ia_i + \tau Ia_i;$$

$$\frac{dR_i}{dt} = \gamma Ia_i - \lambda_i R_i - \delta R_i - \varphi_k R_i + \Upsilon R_i^{v1} + \tau R_i;$$

$$\frac{dS_i^{v1}}{dt} = \delta R_i^{v1} - \lambda_i S_i^{v1} + \varphi_k S_i - \Upsilon S_i^{v1} + \tau S_i^{v1};$$

$$\frac{dE_i^{v1}}{dt} = \lambda_i S_i^{v1} - \sigma_{v1}\varepsilon E_i^{v1} - \varepsilon(1-\sigma_{v1})E_i^{v1} + \tau E_i^{v1};$$

$$\frac{dIs_i^{v1}}{dt} = \sigma_{v1}\varepsilon E_i^{v1} - \psi Is_i^{v1} + \tau Is_i^{v1};$$

$$\frac{dIa_i^{v1}}{dt} = (1-\sigma_{v1})\varepsilon E_i^{v1} + \psi Is_i^{v1} + \lambda_i R_i^{v1} - \gamma Ia_i^{v1} + \tau Ia_i^{v1};$$

$$\frac{dR_i^{v1}}{dt} = \gamma Ia_i^{v1} - \lambda_i R_i^{v1} - \delta R_i^{v1} + \varphi_k R_i - \Upsilon R_i^{v1} + \tau R_i^{v1};$$

$\lambda_i = q_i \kappa \sum_{j=1}^{4} c_{ij}\left(Is_i + \rho(Ia_i) + Is_i^{v1} + \rho(Ia_i^{v1})\right);$ force of infection

$\kappa = 1 + w_1 \cos\left(\frac{2\pi t}{364} + w_2\right)$ seasonal forcing

#### S1.2 Input parameters for transmission model

Table 1: Input parameters for the dynamic transmission model.

| Parameter | Symbol | Input value (mean) | 95% CI | Distribution | Source |
|---|---|---|---|---|---|
| Duration of symptoms (days) | $1/\psi$ | 2 days | - | Fixed value | (1–3) |
| Duration of asymptomatic shedding (days) | $1/\gamma$ | 10 days | - | Fixed value | (1,3,4) |
| Incubation period | $1/\varepsilon$ | 1 day | - | Fixed value | (1–3) |
| Aging | $T$ | 1/365 | - | Fixed value | (5) |
| Birth rate | $1/b$ | 11.4 per 1000 | - | Fixed value | (6) |
| Contact rate | $c_{ij}$ | Supplement Figure 5 | - | Fixed value | (7) |
| Relative infectiousness during asymptomatic period | $\rho$ | 0.05 | - | Fixed value | (1,8) |
| Proportion of infections symptomatic | $\sigma$ | 0.86 | 0.77-0.90 | Beta(94.1, 15.3) | (5) |
| Proportion of infections symptomatic in one dose vaccinated | $\sigma_{v1}$ | 0.6 | 0.3-0.9 | Fixed value | Base case (sensitivity analysis) |
| Duration of immunity (years) | $1/\delta$ | 8.89 | 5.84-11.8 | Gaussian | (5) |
| Seasonal amplitude term | $w_1$ | 0.019 | 0.016-0.021 | Gaussian | (5) |
| Seasonal offset term | $w_2$ | 0.211 | 0.080-0.340 | Gaussian | (5) |
| Probability of infection between under 5s | $q_1$ | 0.18 | 0.15-0.21 | Beta(146.1-665.4) | (5) |
| Probability of infection to over 5s | $q_2$ | 0.04 | 0.037-0.047 | Beta(247.9-5793.8) | (5) |
| Vaccination rate (coverage) <5 | $\varphi_{0\text{-}4}$ | 90% | - | Fixed value | (9) |
| Vaccination rate (coverage) 65+ | $\varphi_{65+}$ | 75% | - | Fixed value | (10) |
| Vaccination waning dose 1 | $\Upsilon$ | 2 years | - | (6mo – 8.9 yrs) | Base case (sensitivity analysis) |

#### S1.3 Additional model details

Births only flow into the 0-4 age group. Death is represented by aging out of the oldest age group. No additional age specific death rate was implemented. Aging was implemented as described by King and Wearing et al. (11). Probability of transmission was represented as a 2 x 2 matrix:

| | Contact under 5 years old | Contact over 5 years old |
|---|---|---|
| Infected under 5 years old | *q1* | *q2* |
| Infected over 5 years old | *q2* | *q2* |

Where *q1* represent the probability of transmission from an under 5 years old infected child to an under 5 years old contact. Conversely, *q2* represents the probability of transmission from an under 5 years old infected child to an over 5 years old contact. The same probability of *q2* is applied for transmission from an over 5 years old infected individual to an under 5 years old child, and an over 5 years old infected individual to an over 5 years old contact. Further details of the transmission model structure and features are described in Bolt et al. (5).

Contact is represented with $c_{ij}$ with the following UK contact matrix from POLYMOD (7):

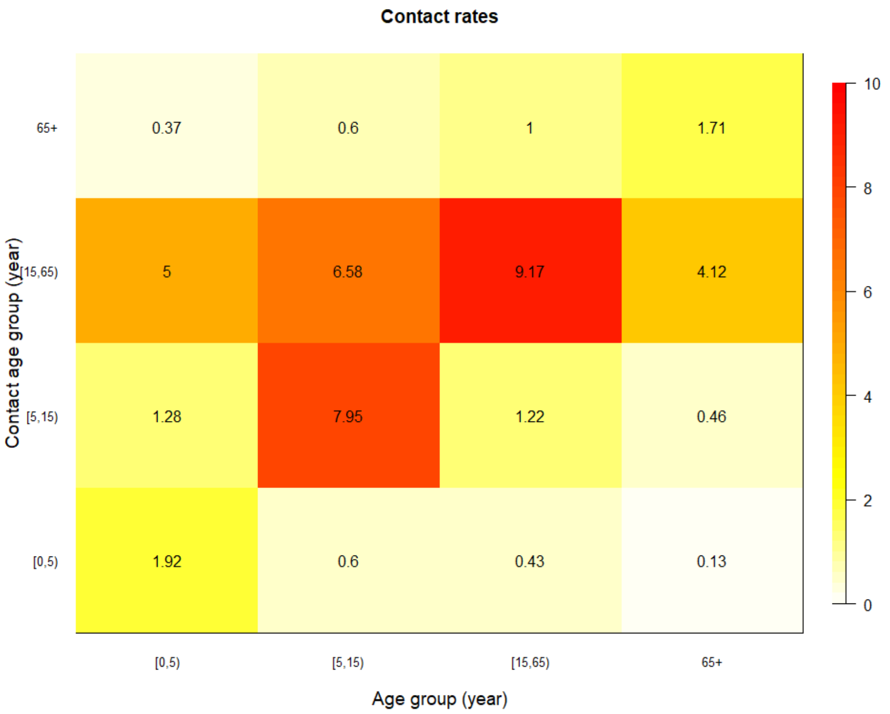

Force of infection is contributed by those who were symptomatic unvaccinated, asymptomatic unvaccinated, symptomatic vaccinated, and asymptomatic unvaccinated. We assume that unvaccinated asymptomatic individuals are less infectious by a factor ρ. If individuals were vaccinated and infected they were less likely to be symptomatic by a factor $\sigma_{v1}$, and more likely to be asymptomatic by a factor 1- $\sigma_{v1}$. Vaccination rate was implemented as:

$$vaccination\ rate = \frac{target\ coverage}{numebr\ of\ days\ to\ reach\ coveage}$$

where the number of days to reach coverage was assumed to be within 365 days.

#### S1.4 Input parameters for health outcomes

Table 2: Input parameters for probability of health outcomes of GP attendance, hospitalisation due to norovirus, hospitalisation due to norovirus induced AKI, mortality due to norovirus, and mortality due to norovirus induced AKI, stratified by age.

| Parameter | Input value (mean) | 95% CI | Distribution | Source |
|---|---|---|---|---|
| Norovirus GP attendance | | | | |
| 0-4 | 0.046 | 0.043-0.049 | Beta(389.9, 8377.2) | (5) |
| 5-14 | 0.044 | 0.040-0.049 | Beta(389.9, 8377.2) | (5) |
| 15-64 | 0.0011 | 0.0007-0.0018 | Beta(15.3, 13937.7) | (12) |
| 65+ | 0.0021 | 0.0011-0.004 | Beta(8, 3819.9) | (12) |
| Norovirus hospitalisation (annual) | | | | |
| 0-4 | 0.0033 | 0.00241-0.0044 | Beta(40.5, 12223.3) | (13) |
| 5-14 | 0.0004 | 0.0003-0.0005 | Beta(61.4, 153540.1) | (13) |
| 15-64 | 0.00026 | 0.0002-0.00033 | Beta(61.4, 236282.2) | (13) |
| 65+ | 0.037 | 0.027-0.048 | Beta(44.4, 1143.3) | (5) |
| Acute kidney injury hospitalisation (annual) | | | | |
| 0-4 | - | | - | - |
| 5-14 | - | | - | - |
| 15-64 | - | | - | - |
| 65+ | 0.126 | 0.075-0.180 | Beta(19.2, 133.4) | (5) |
| Norovirus mortality | | | | |
| 0-4 | 0.00000625 | 0.0000057-0.0000067 | Beta(600.2, 96038798.5) | (14) |
| 5-14 | 0.00000466 | 0.0000043–0.000005 | Beta(681, 146136237) | (14) |
| 15-64 | 0.00000466 | 0.0000043–0.000005 | Beta(681, 146136237) | (14) |
| 65+ | 0.000435 | 0.0004–0.00047 | Beta(593.2, 1362972.4) | (14) |
| Acute kidney injury mortality | | | | |
| 0-4† | 0.033 | 0.02-0.12 | Beta(1.7, 49.4) | (15) |
| 5-14† | 0.033 | 0.02-0.12 | Beta(1.7, 49.4) | (15) |
| 15-64‡ | 0.0685 | 0.014-0.254 | Beta(1.1, 14.9) | (15) |
| 65+¥ | 0.222 | 0.116-0.45 | Beta(5.1, 17.7) | (15) |

† Unadjusted mortality age <18 years old, range from AKI stage 1 (0.02) to AKI stage 3 (0.18)

‡ Unadjusted mortality age18-64 years old, range from AKI stage 1 of 18-39 year olds (0.014) to AKI stage 3 of 40-64 year olds  (0.254)

¥ Unadjusted mortality age 65+ years old, range from AKI stage 1 of 65-74 year olds (0.12) to AKI stage 3 of 75+ year olds  (0.45)

#### S1.5 Input parameters quality-of-life-loss per episode

Table 3: Input parameter for quality-of-life loss per episode of GP attendance, hospitalisation due to norovirus, hospitalisation due to norovirus induced AKI, stratified by age.

| Parameter | | QALY loss per episode | Range | Distribution | Source |
|---|---|---|---|---|---|
| Norovirus GP attendance* | | | | | |
| | 0-4 | 0.00257 | 0.00144-0.00367 | Beta(20.3, 7900.9) | (16) |
| | 5-14 | 0.00257 | 0.00144-0.00367 | Beta(20.3, 7900.9) | (16) |
| | 15-64 | 0.00257 | 0.00144-0.00367 | Beta(20.3, 7900.9) | (16) |
| | 65+ | 0.00257 | 0.00144-0.00367 | Beta(20.3, 7900.9) | (16) |
| Norovirus hospitalisation† | | | | | |
| | 0-4 | 0.00715 | 0.00565-0.00867 | Beta(85.9, 11925.3) | (16) |
| | 5-14 | 0.00715 | 0.00565-0.00867 | Beta(85.9, 11925.3) | (16) |
| | 15-64 | 0.00715 | 0.00565-0.00867 | Beta(85.9, 11925.3) | (16) |
| | 65+ | 0.00715 | 0.00565-0.00867 | Beta(85.9, 11925.3) | (16) |
| Acute kidney injury hospitalisation‡ | | | | | |
| | 0-4 | 0.00493 | 0-0.0096 | Beta(4, 813.2) | (17) |
| | 5-14 | 0.00493 | 0-0.0096 | Beta(4, 813.2) | (17) |
| | 15-64 | 0.00493 | 0-0.0096 | Beta(4, 813.2) | (17) |
| | 65+ | 0.00493 | 0-0.0096 | Beta(4, 813.2) | (17) |

* Based on general practitioner health utility score of 0.688 (0.553-0.824) for age 18 months to 5 years. Duration of an episode was assumed to be 3 days.

† Based on paediatrician utility score of 0.542 (0.445-0.638) for age 18 months to 5 years. Duration of episode was assumed to be 5.7 days.

‡ AKI patient health utility score of 0.676 (0.520-1.00) for all AKI stages for mean age 65 (IQR 53-74). Duration of episode was assumed to be 12 days.

#### S1.6 Input parameters costs

Table 4: Input parameters for costs of GP attendance, hospitalisation due to norovirus, hospitalisation due to norovirus induced AKI, and vaccine administration cost stratified by age. Costs inflated by 2% per year over the time horizon.

| Parameter | | Cost value (£) | Range | SD | Distribution | Source |
|---|---|---|---|---|---|---|
| Norovirus GP attendance | | | | | | |
| | 0-4 | 49 | 36.75-61.25 | 6.41 | Lognormal | (18) |
| | 5-14 | 49 | 36.75-61.25 | 6.41 | Lognormal | (18) |
| | 15-64 | 49 | 36.75-61.25 | 6.41 | Lognormal | (18) |
| | 65+ | 49 | 36.75-61.25 | 6.41 | Lognormal | (18) |
| Norovirus hospitalisation* | | | | | | |
| | 0-4 | 1045 | 543-3600 | 535 | Lognormal | (19) |
| | 5-14 | 1045 | 543-3600 | 535 | Lognormal | (19) |
| | 15-64 | 1749 | 403-8855 | 1623 | Lognormal | (19) |
| | 65+ | 1749 | 403-8855 | 1623 | Lognormal | (19) |
| Acute kidney injury hospitalisation* | | | | | | |
| | 0-4 | - | - | - | Lognormal | (19) |
| | 5-14 | - | - | - | Lognormal | (19) |
| | 15-64 | - | - | - | Lognormal | (19) |
| | 65+ | 3730 | 2434-10787 | 2131 | Lognormal | (19) |
| Vaccine administration cost* | | | | | | |
| | 0-14 | 18.20 | 9.71-25.97 | 4.64 | Lognormal | (20) |
| | 15+ | 14.05 | 7.59-20.88 | 3.69 | Lognormal | (20) |

* Costs inflated to 2022/2023 values using the NHS cost inflation index (18)

### S2. Additional results

#### S2.1 Sampling distributions

Latin hypercube sampled parameters (n = 2000)

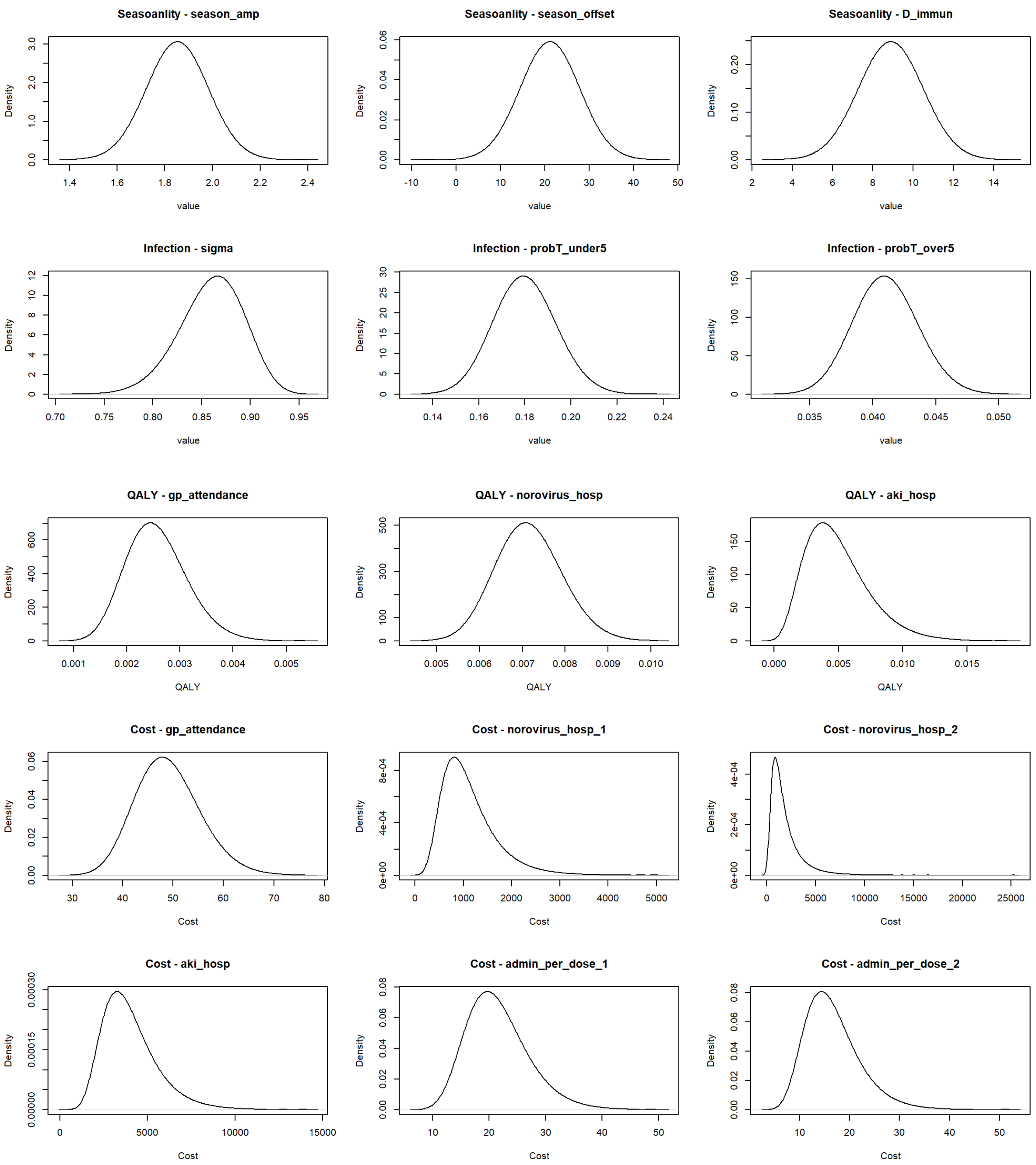

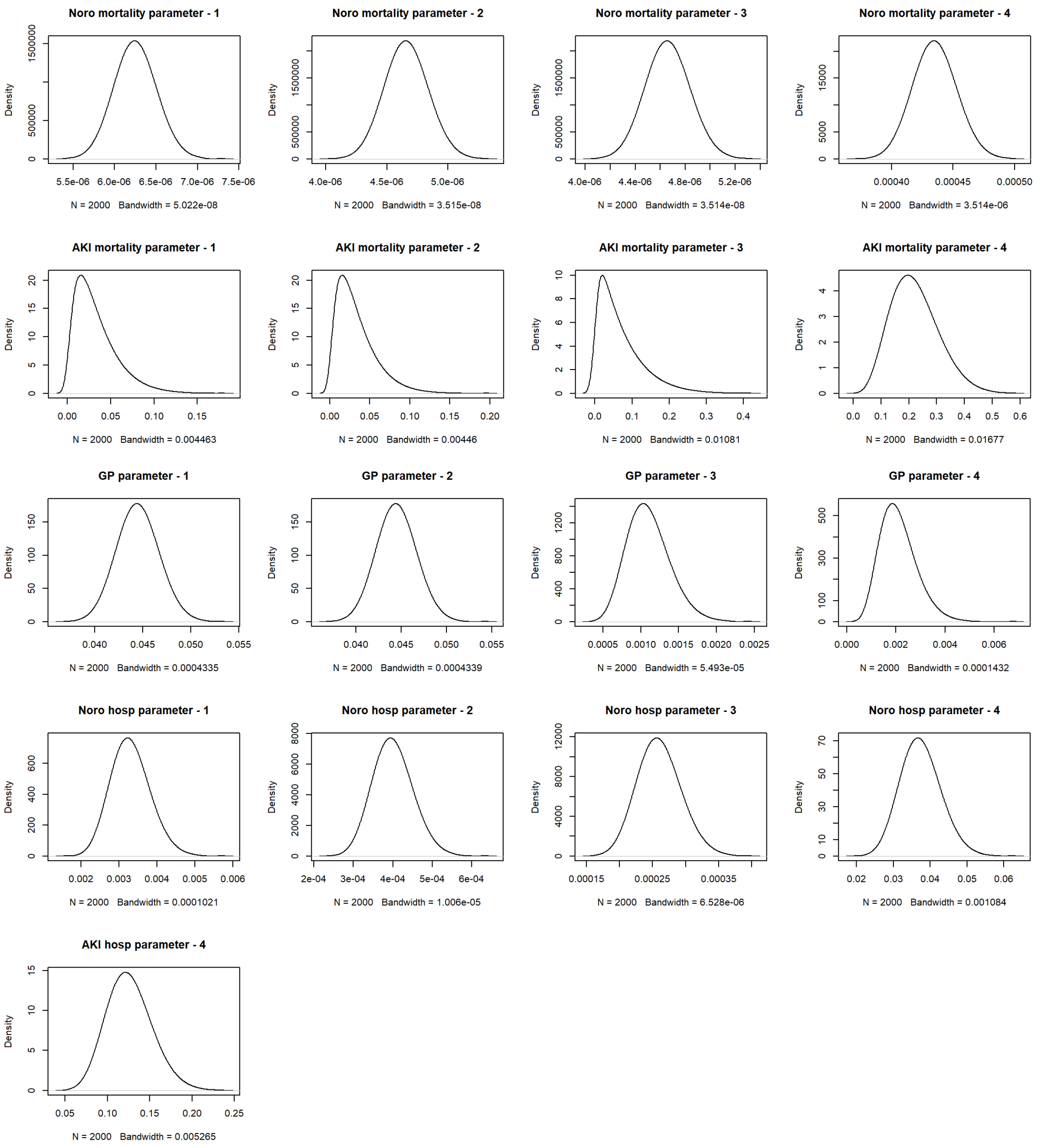

Noro mortality parameter - 1
N = 2000 Bandwidth = 5.022e-08
Noro mortality parameter - 2
N = 2000 Bandwidth = 3.515e-08
Noro mortality parameter - 3
N = 2000 Bandwidth = 3.514e-08
Noro mortality parameter - 4
N = 2000 Bandwidth = 3.514e-06
AKI mortality parameter - 1
N = 2000 Bandwidth = 0.004463
AKI mortality parameter - 2
N = 2000 Bandwidth = 0.00446
AKI mortality parameter - 3
N = 2000 Bandwidth = 0.01081
AKI mortality parameter - 4
N = 2000 Bandwidth = 0.01677
GP parameter - 1
N = 2000 Bandwidth = 0.0004335
GP parameter - 2
N = 2000 Bandwidth = 0.0004339
GP parameter - 3
N = 2000 Bandwidth = 5.493e-05
GP parameter - 4
N = 2000 Bandwidth = 0.0001432
Noro hosp parameter - 1
N = 2000 Bandwidth = 0.0001021
Noro hosp parameter - 2
N = 2000 Bandwidth = 1.006e-05
Noro hosp parameter - 3
N = 2000 Bandwidth = 6.528e-06
Noro hosp parameter - 4
N = 2000 Bandwidth = 0.001084
AKI hosp parameter - 4
N = 2000 Bandwidth = 0.005265
Density

#### S2.2 Incidence of symptomatic cases averted

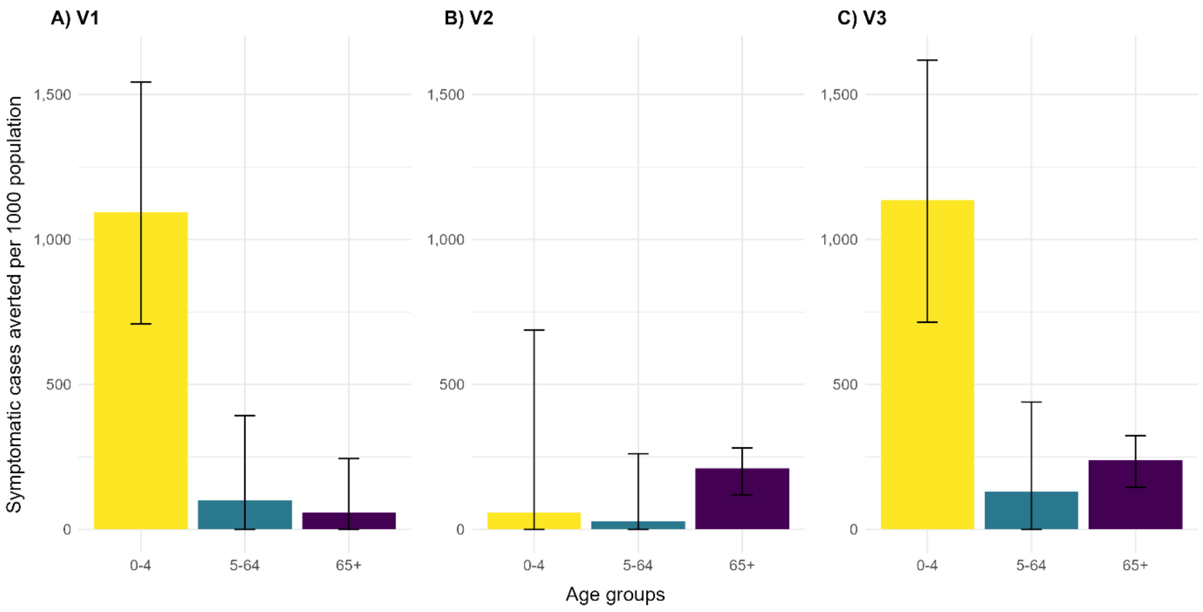

Figure 1: Incidence per 1000 population of symptomatic norovirus cases averted per age group by vaccination scenarios in A) V1 B) V2 C) V3. Yellow bars represent reduction in incidence in children aged 0-4. Blue bars represent reduction in incidence in individuals aged 5-64. Purple bars represent reduction in incidence in adults aged 65+.

### S2.3 Quality of life gains by vaccination strategy and discounting (including AKI outcomes)

Table 5: Discounted quality of life gains by vaccination strategy (including AKI outcomes), age group, and health outcome

| Vaccination strategy | Age group | GP attendance | Norovirus hospitalisation | Norovirus mortality | AKI hospitalisation | AKI mortality |
|---|---|---|---|---|---|---|
| **V1 under 5** | | | | | | |
| | 0-4 | 422 (246-653) | 87 (56-126) | 530 (418-620) | 0 (0-0) | 0 (0-0) |
| | 15-64 | 10 (4-19) | 7 (4-10) | 321 (239-429) | 0 (0-0) | 0 (0-0) |
| | 5-14 | 125 (74-195) | 3 (2-5) | 115 (98-143) | 0 (0-0) | 0 (0-0) |
| | 65+ | 4 (1-7) | 180 (116-269) | 1,925 (1,561-2,399) | 417 (100-1,016) | 123,962 (39,599-260,635) |
| **V2 over 65** | | | | | | |
| | 0-4 | 24 (14-37) | 5 (3-7) | 30 (23-36) | 0 (0-0) | 0 (0-0) |
| | 15-64 | 3 (1-6) | 2 (1-3) | 98 (79-123) | 0 (0-0) | 0 (0-0) |
| | 5-14 | 22 (13-36) | 1 (0-1) | 20 (16-27) | 0 (0-0) | 0 (0-0) |
| | 65+ | 12 (4-27) | 604 (324-987) | 6,485 (3,831-9,486) | 1,399 (328-3,423) | 415,040 (119,180-917,897) |
| **V3 under 5 and over 65** | | | | | | |
| | 0-4 | 439 (257-678) | 91 (58-131) | 551 (439-642) | 0 (0-0) | 0 (0-0) |
| | 15-64 | 13 (6-25) | 9 (6-13) | 427 (340-545) | 0 (0-0) | 0 (0-0) |
| | 5-14 | 149 (88-233) | 4 (3-5) | 138 (116-172) | 0 (0-0) | 0 (0-0) |
| | 65+ | 14 (5-30) | 692 (397-1,102) | 7,424 (4,913-10,387) | 1,602 (381-3,860) | 475,522 (142,076-1,036,239) |

Table 6: Un-discounted quality of life gains by vaccination strategy (including AKI outcomes), age group, and health outcome

| Vaccination strategy | Age group | GP attendance | Norovirus hospitalisation | Norovirus mortality | AKI hospitalisation | AKI mortality |
|---|---|---|---|---|---|---|
| **V1 under 5** | | | | | | |
| | 0-4 | 482 (280-746) | 100 (63-144) | 605 (476-708) | 0 (0-0) | 0 (0-0) |
| | 15-64 | 11 (5-21) | 7 (5-11) | 349 (258-476) | 0 (0-0) | 0 (0-0) |
| | 5-14 | 139 (83-218) | 3 (2-5) | 128 (108-160) | 0 (0-0) | 0 (0-0) |
| | 65+ | 4 (1-8) | 196 (125-296) | 2,098 (1,675-2,667) | 454 (108-1,111) | 135,089 (43,164-284,159) |
| **V2 over 65** | | | | | | |
| | 0-4 | 26 (15-40) | 5 (3-8) | 32 (25-40) | 0 (0-0) | 0 (0-0) |
| | 15-64 | 3 (2-6) | 2 (1-3) | 108 (86-136) | 0 (0-0) | 0 (0-0) |
| | 5-14 | 24 (14-39) | 1 (0-1) | 22 (16-30) | 0 (0-0) | 0 (0-0) |
| | 65+ | 14 (5-31) | 695 (372-1,137) | 7,461 (4,407-10,909) | 1,609 (379-3,943) | 477,487 (137,210-1,055,052) |
| **V3 under 5 and over 65** | | | | | | |
| | 0-4 | 500 (292-773) | 103 (66-149) | 628 (500-731) | 0 (0-0) | 0 (0-0) |
| | 15-64 | 15 (6-27) | 10 (6-14) | 465 (365-604) | 0 (0-0) | 0 (0-0) |
| | 5-14 | 165 (98-258) | 4 (3-6) | 152 (127-191) | 0 (0-0) | 0 (0-0) |
| | 65+ | 16 (5-34) | 790 (452-1,258) | 8,474 (5,604-11,906) | 1,828 (434-4,406) | 542,698 (161,659-1,179,538) |

#### S2.4 Costs by strategy and discounting (including AKI outcomes)

Table 7: Discounted total costs (GBP) of health care outcomes by vaccination strategy (including AKI outcomes), age group, and health outcome

| Vaccination strategy | Age group | GP attendance cost | Norovirus hospitalisation cost | AKI hospitalisation cost |
|---|---|---|---|---|
| **V1 under 5** | | | | |
| | 0-4 | 8,768,667 (6,182,116-11,892,886) | 14,887,182 (4,691,268-35,115,806) | 0 (0-0) |
| | 15-64 | 203,383 (100,979-357,870) | 1,829,606 (268,518-6,316,629) | 0 (0-0) |
| | 5-14 | 2,556,146 (1,860,572-3,530,897) | 525,772 (171,428-1,287,618) | 0 (0-0) |
| | 65+ | 73,295 (29,591-139,851) | 49,849,559 (7,179,167-175,109,080) | 357,845,354 (131,902,693-760,936,024) |
| **V2 over 65** | | | | |
| | 0-4 | 476,452 (333,124-667,262) | 809,502 (255,985-1,951,984) | 0 (0-0) |
| | 15-64 | 62,692 (32,400-107,646) | 568,305 (84,388-1,909,762) | 0 (0-0) |
| | 5-14 | 444,420 (302,628-649,220) | 91,522 (28,702-228,799) | 0 (0-0) |
| | 65+ | 254,505 (88,522-519,144) | 175,585,726 (24,295,535-593,523,283) | 1,249,115,740 (400,666,048-2,791,047,140) |
| **V3 under 5 and over 65** | | | | |
| | 0-4 | 9,105,993 (6,498,333-12,309,661) | 15,459,141 (4,885,866-36,434,277) | 0 (0-0) |
| | 15-64 | 270,941 (137,923-467,242) | 2,440,415 (361,347-8,203,039) | 0 (0-0) |
| | 5-14 | 3,044,462 (2,194,617-4,227,571) | 626,152 (205,436-1,526,264) | 0 (0-0) |
| | 65+ | 290,061 (106,424-583,286) | 199,780,863 (27,682,922-665,137,464) | 1,422,722,301 (483,542,428-3,106,721,600) |

Table 8: Un-discounted total costs (GBP) of health care outcomes by vaccination strategy (including AKI outcomes), age group, and health outcome

| Vaccination strategy | Age group | GP attendance cost | Norovirus hospitalisation cost | AKI hospitalisation cost |
|---|---|---|---|---|
| **V1 under 5** | | | | |
| | 0-4 | 10,057,131 (7,088,194-13,663,255) | 17,075,000 (5,383,965-40,291,212) | 0 (0-0) |
| | 15-64 | 222,482 (109,913-393,412) | 2,000,933 (288,187-6,940,020) | 0 (0-0) |
| | 5-14 | 2,849,879 (2,066,603-3,977,477) | 586,274 (191,799-1,442,270) | 0 (0-0) |
| | 65+ | 80,322 (32,285-153,413) | 54,626,837 (7,833,979-190,435,609) | 392,178,282 (144,771,697-838,356,202) |
| **V2 over 65** | | | | |
| | 0-4 | 520,953 (361,841-733,236) | 885,206 (278,177-2,148,596) | 0 (0-0) |
| | 15-64 | 69,197 (35,757-118,774) | 627,193 (93,576-2,119,710) | 0 (0-0) |
| | 5-14 | 482,062 (325,158-714,296) | 99,288 (30,618-247,258) | 0 (0-0) |
| | 65+ | 294,316 (102,323-601,316) | 203,049,949 (28,095,643-685,724,213) | 1,444,519,479 (463,240,192-3,227,622,191) |
| **V3 under 5 and over 65** | | | | |
| | 0-4 | 10,424,114 (7,419,514-14,113,304) | 17,697,043 (5,603,122-41,707,824) | 0 (0-0) |
| | 15-64 | 296,943 (150,955-513,901) | 2,673,571 (393,811-8,974,941) | 0 (0-0) |
| | 5-14 | 3,376,651 (2,429,450-4,716,912) | 694,532 (228,390-1,703,423) | 0 (0-0) |
| | 65+ | 332,853 (121,888-670,489) | 229,261,749 (31,772,190-761,518,911) | 1,632,662,132 (553,552,868-3,566,750,100) |

### S2.5 One way sensitivity analysis (AKI outcomes excluded)

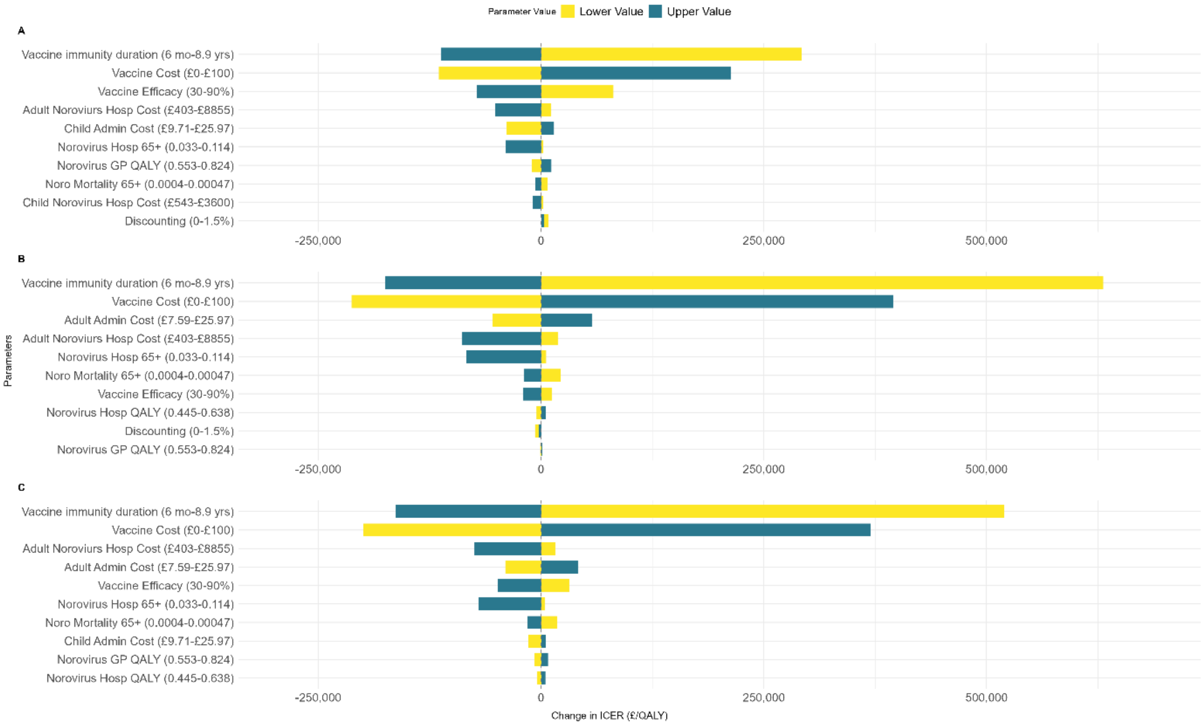

Figure 3: One way sensitivity analysis by vaccination strategy without AKI outcomes A) V1; average ICER £164,054 per QALY B) V2; average ICER £289,398 per QALY C) V3; average ICER £276,572 per QALY as reported in Table 2. Average ICERs reported vs baseline.

#### S2.6 Vaccination cost threshold analysis

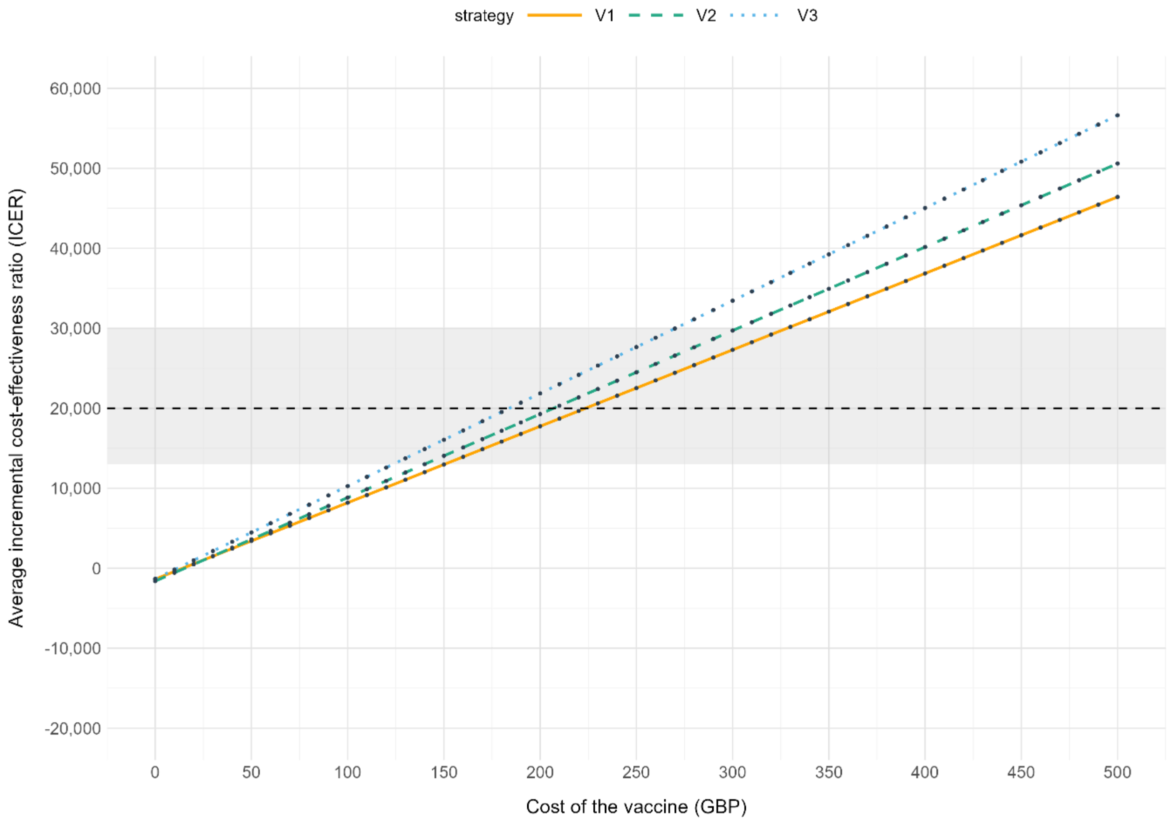

Figure 4: Threshold analysis of the cost of the vaccine (GBP). Plots of average ICERs vs baseline by vaccination strategy V1, V2, V3. Dashed line indicates £20,000 per QALY ICER. Shaded area indicates ICER £13,000 - £30,000 per QALY.

### S2.7 Incremental net monetary benefit (INMB)

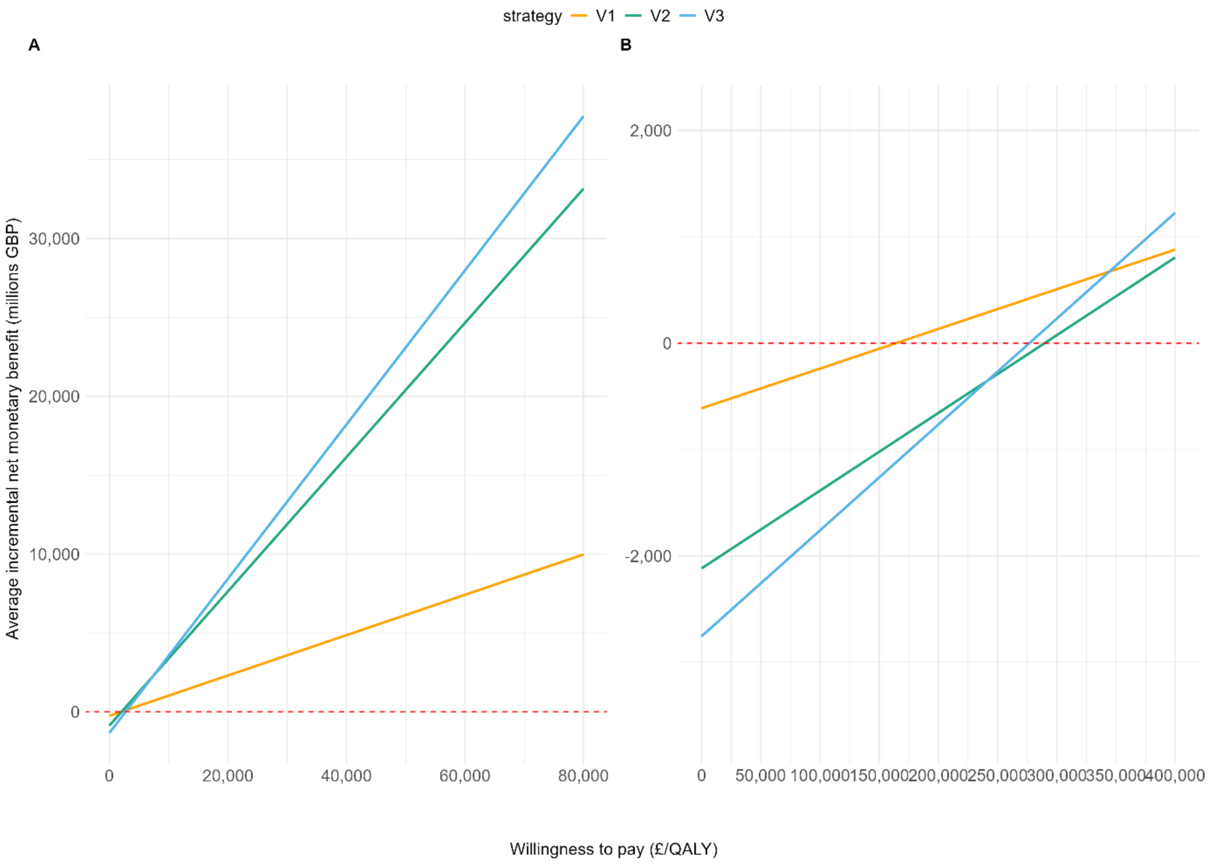

Figure 5: Incremental net monetary benefit by vaccination strategy vs baseline by A) including AKI outcomes, B) excluding AKI outcomes. Dashed line indicates 0 net monetary benefit. Colours denote vaccination strategy: orange is V1, green is V2, and blue is V3.
